# Modeling the Effective Control Strategy for the Transmission Dynamics of Global Pandemic COVID-19

**DOI:** 10.1101/2020.04.22.20076158

**Authors:** M. H. A. Biswas, M. S. Khatun, A. K. Paul, M. R. Khatun, M. A. Islam, S. A. Samad, U. Ghosh

**Affiliations:** Mathematics Discipline, Khulna University, Khulna, Bangladesh; Department of Applied Mathematics, University of Calcutta, Kolkata, WB, India

**Keywords:** COVID-19, SARS CoV-2, Mathematical model, Maximum principle, Optimal control, Social distancing

## Abstract

The novel coronavirus disease (namely COVID-19) has taken attention because of its deadliness across the globe, causing a massive death as well as critical situation around the world. It is an infectious disease which is caused by newly discovered coronavirus. Our study demonstrates with a nonlinear model of this devastating COVID-19 which narrates transmission from human-to-human in the society. Pontryagin’s Maximum principle has also been applied in order to obtain optimal control strategies where the maintenance of social distancing is the major control. The target of this study is to find out the most fruitful control measures of averting coronavirus infection and eventually, curtailed of the COVID-19 transmission among people. The model is investigated analytically by using most familiar necessary conditions of Pontryagin’s maximum principle. Furthermore, numerical simulations have been performed to illustrate the analytical results. The analysis reveals that implementation of educational campaign, social distancing and developing human immune system are the major factors which can be able to plunge the scenario of becoming infected.

## 1. Introduction

The recent outbreak of COVID-19 (coronavirus disease) has newly emerged at the end of 2019 in Wuhan (capital of Hubei province, China), which has now become a global pandemic. In 2003, Severe Acute Respiratory Syndrome (SARS) Coronavirus first identified whereas first infected humans were detected in the Guangdong province of Southern China in 2002. Almost 26 countries affected by this SARS-CoV causing above 8000 cases in 2003 [24]. The newly emerged COVID-19 caused by coronavirus is genetically identical to SARS-CoV 2002. Hence, the scientists have named this virus as Severe Acute Respiratory Syndrome Coronavirus-2 (SARS CoV-2). It is thought that SARS-CoV is an animal virus from an animal reservoir, probably bats that spread to other animal like civet cats. COVID-19 is a human-to-human transmittable disease. It seems to have occurred mainly during the second week of illness. The symptoms of this disease include fever, malaise, myalgia, cough, shortness of breath, diarrhoea, headache and shivering. There is no specific symptom for the diagnosis of SARS. Though fever is the most frequent symptom but in adult and immunosuppressed patients it is most often absent on initial stage.

In 2020 on 2 April, there were approximately 81,589 cases of coronavirus in China. Among which about 76,408 people got recovered whereas almost 3,318 people died from this devastating disease [11]. But the wonderful matter is that the currently infected populations have been decreased because of the implementation of proper measures and consciousness among people in China. Besides, on 13 March, World Health Organization declared Europe as the most epicenter of COVID-19 the pandemic disease. Among European countries, Italy has the most cases outside of China followed by Spain, Germany, France, Switzerland, Netherlands, Austria, and Belgium. But now the scenario has completely been changed. At present United States has become the top spot in the case of COVID-19 related incidence and death in the world.

There is an extensive body of work to formulate mathematical models and optimal control strategies of emerging infectious diseases. Biswas *et al*. [1] developed a SEIR model for infectious diseases and also employed optimal control strategy introducing state constraints on the model. In this continuation, Biswas *et al*. [9] proposed a nonlinear model to show the optimal immunotherapeutic treatment for HIV. See also Biswas [2-8] where he showed mastery knowledge in investigating and analyzing the mechanisms of the most devastating diseases in which mathematical modeling and optimal control technique acted as the key tool. Several well documented models of COVID-19 have been presented as well. Kucharski *et al*. [17] studied a mathematical model on early dynamics of transmission and control of COVID-19. Zhuang *et al*. [31] estimated the COVID 19 cases by the imported cases and air travel data from Iran to other Middle East countries. Zhang *et al*. [30] presented a data-driven analysis on the estimation on the reproductive number of COVID-19 and the probable outbreak size on the Diamond Princess cruise ship. A modeling study on domestic and international spread of COVID-19 has been analyzed by in detail Wu *et al*. [25]. Lin *et al*. [21] proposed a conceptual model for the COVID-19 outbreak in Wuhan with the consideration of individual behavioral reaction and governmental actions. Zhang *et al*. [29] developed a new psychological crisis intervention model by utilizing internet technology. Khan and Atangana [18] modeled the dynamics of novel coronavirus with fractional derivative. They applied the Atangana-Baleanu derivative because of their kernel being nonlocal and nonsingular and also for their crossover behavior. An accurate early forecasting model from small dataset regarding nCOVID-19 has been analyzed by Fong *et al*. [15]. Readers can also follow (Zhao *et al*. [27]; Zhao *et al*. [28]; Sookaromdee and Wiwanitkit [23]; Chen *et al*. [14]; Chen and Yu [13]; Xu *et al*. [26]; Khatun and Biswas [19, 20] and references within) for more detailed information about novel coronavirus (COVID-19) as well as very recent implementation of modeling and optimal control techniques.

In view of the gravity that associates with decisions to be made about the pandemic COVID-19, there needs an urgent and serious coordinated response from all sectors to combat against the highly infectious disease as soon as possible. As part of this coordinated approach, we present a mathematical model of novel coronavirus (COVID-19) to show the transmission which can be controlled effectively by maintaining social distancing as well as treatment. In this study, we analyze the mathematical modeling of COVID-19 and also apply optimal control theory introducing three controls to prevent the transmission of this severe infectious disease. Finally we perform numerical simulations to interpret the outcomes and also to show the effectiveness of the controls in preventing and/or curing the infection. The main aim of this work is to minimize the infection by applying the controls and also the associated cost of the implementation of the three controls measures.

## 2. Current Scenario of COVID-19

At present (on 02 April, 2020) USA has the most COVID-19 cases outside of China (about 81,589 infections) with about 215,344 incidences, followed by Italy at 110,574, Spain at 110,238, Germany at 77,981, France at 56, 989, Iran at 47,593 and other countries. Besides, other parts in Europe the cases have been becoming soared nowadays. As for example, currently United Kingdom has 29,474 incidences, followed by Switzerland at 17,781, Belgium at 15,348, Netherlands at 13,614, and Austria at 10,842. But it is noticeable that the death rate in USA is lower than Italy. The COVID 19 related death rate is very high in Italy (at 13,155) followed by Spain at 10,003, France at 4,032 and the COVID 19 related recovery rate is raised in China (at about 76,408) followed by Spain at 26,743, Italy at 16,847. The detailed about coronavirus related cases, deaths from the disease and recovered rate are presented in Figure 1 [10]. The novel coronavirus-2 disease (COVID 19) is mysterious because it transmits only through eyes, nose and mouth of the human being and directly targets our lunge. Initially, COVID 19 emerged in China and from there the devastating disease spread out almost 206 countries in the world. Present scenario of COVID 19 related incidence and death rate are shown in Figure 2 [11]. The deadly coronavirus has also broken out in Bangladesh along with other countries in the world. The first coronavirus cases in Bangladesh have been noticed on 8 March 2020 and since then it has been spread out across the country. Now this disease has become an epidemic in Bangladesh. At the beginning of infection in Bangladesh only three persons were affected by this novel coronavirus but the infection has risen considerably. On 16 March, there were about five persons infected from COVID 19 but it had increased on 23 March by nine persons. By this time, about 6 patients have died from this fatal disease COVID 19. However, the biggest problem here is that the new incidence rate is increasing day by day. The present status of COVID 19 cases from March 8 to April 3 are exhibited in Figure 3 [12].

**Figure 1.**
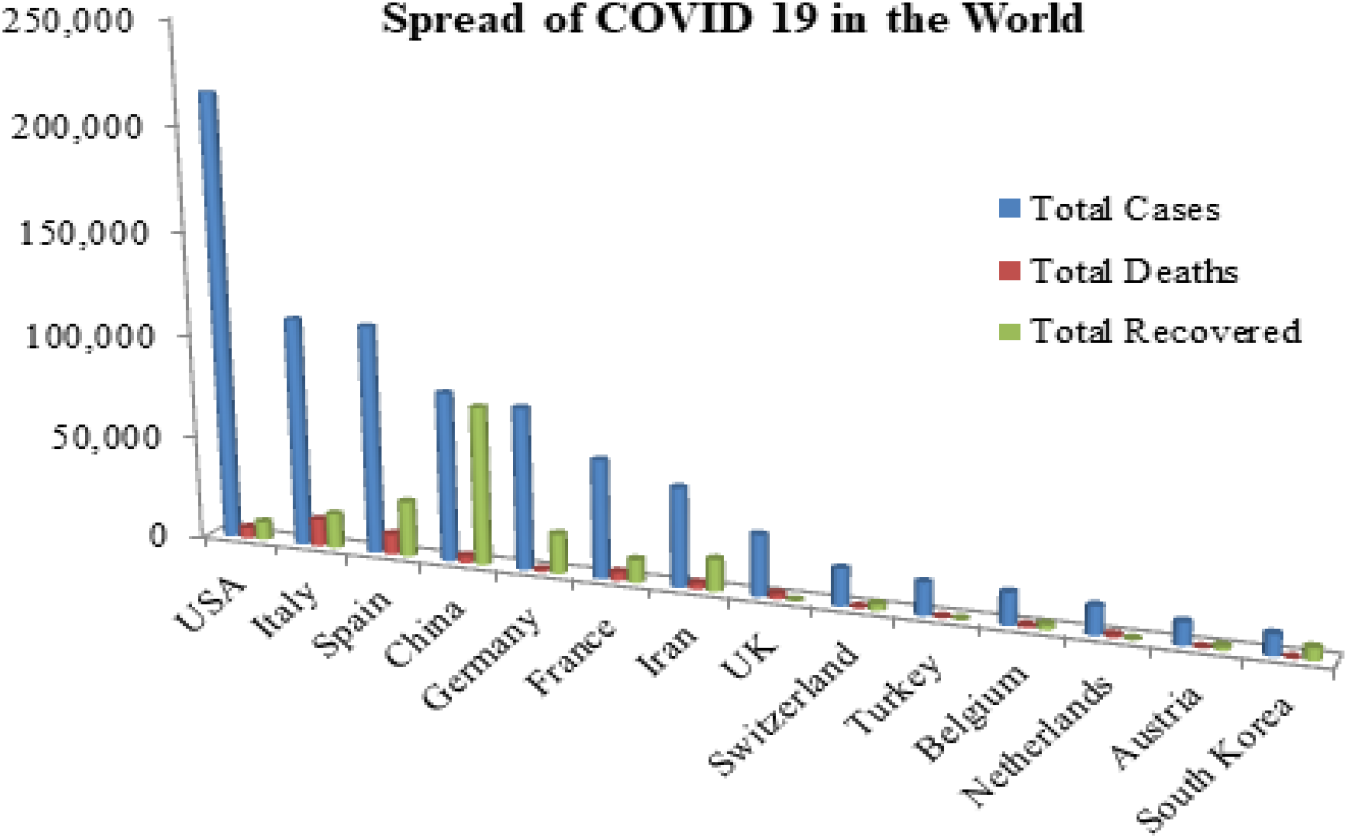
Number of COVID 19 cases, deaths rate and recovery rate in the World (Source: [10]).

**Figure 2.**
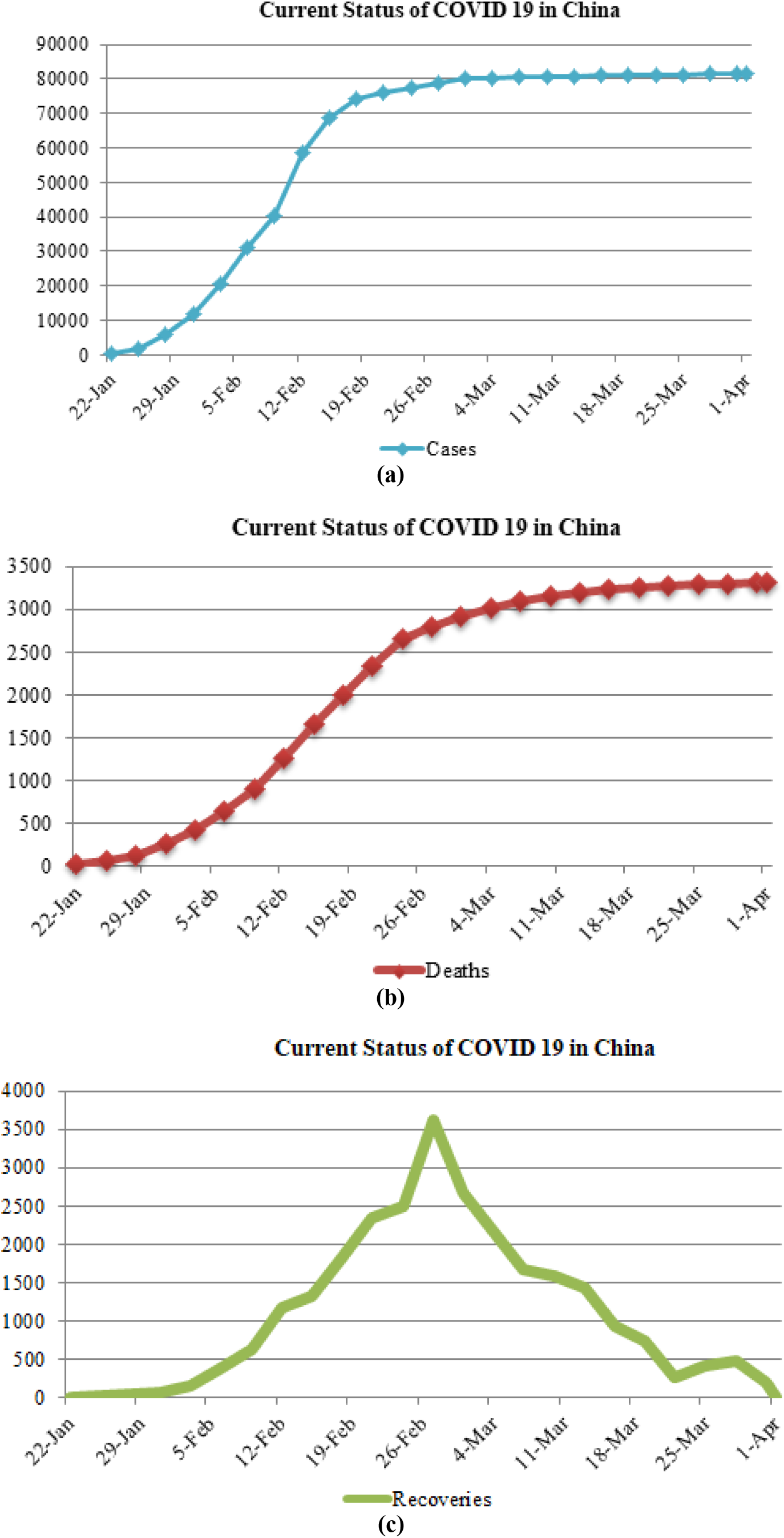
(a) Number COVID 19 cases from January 22 to April 2; (b) Number of COVID 19 related deaths rate from January 22 to April 2; and (c) Number of COVID 19 related recovery rate from January 22 to April 2 (Source: [11]).

**Figure 3.**
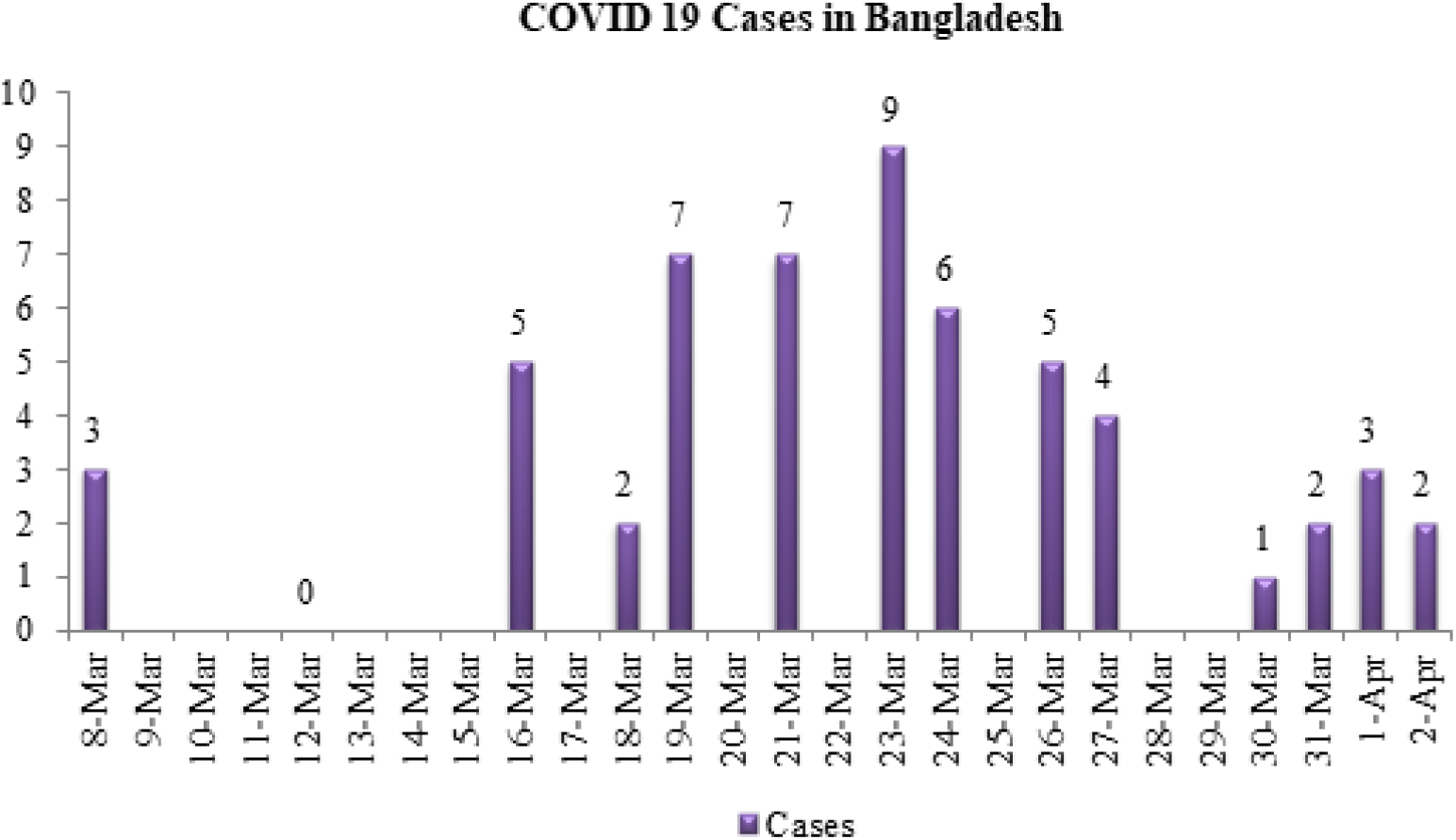
Number of COVID 19 cases in Bangladesh from March 08 to April 2 (Source: [12]).

## 3. Compartmental Model

SARS Coronavirus-2 is a newly discovered mysterious pathogen and there are no appropriate drugs and/or vaccines yet for its treatments. So, in order to find out effective methods for prevention and control of SARS Coronavirus-2 disease, it is indispensable for the physicians and biologists to understand the disease mechanisms in the human. However, SARS coronavirus-2 is such type of virus which first transmitted from animal to human. When it has been transmitted to human, then it continues to be transmitted through human to human by the closed contact of infected individuals due to its highly infectiveness. In the present study we highlight only human to human transmission. So, the whole dynamics of SARS Coronavirus-2 disease (COVID-19) can be described by a SIRS type infectious disease model in terms of a set of nonlinear ordinary differential equations (NODEs). Let us consider that *S*(*t*), *Q*(*t*), *I*(*t*), *I*_*s*_ (*t*), and *R*(*t*) be the number of susceptible, quarantined, infected, isolated and removed individuals respectively at time *t*. Let *N*(*t*) be the total population at time *t* where *N*(*t*) = *S*(*t*) + *Q*(*t*) + *I*(*t*) + *I*_*s*_ (*t*) + *R*(*t*). Here, the susceptible individuals (*S*(*t*)) are those who are not affected by the coronavirus infections but any time they may be infected. The individuals who come from overseas mainly from the COVID-19 affected areas and people who come in contact with those individuals have high possibility of carrying the infection. Therefore, such populations must maintain social distancing to combat the transmission of this highly infectious disease. These populations are denoted by quarantined population, *Q*(*t*) in the model. The Quarantine is actually a restriction on the movement of people who are supposed to be exposed to infectious disease to prevent the spread of the disease. They may have been exposed to the disease but do not know or they may have the disease but do not show symptoms. Such populations are categorized as the quarantined individuals in the present model. As COVID-19 has an incubation period of 2-14 days, the individuals are to be quarantined for this time period until their medical checkup is confirmed. After performing the test, people who are identified as COVID-19 positive are taken as the infected individual, *I*(*t*) in the model and these individuals have that ability to transmit. Individuals who have had the coronavirus infections (COVID-19) and they can transmit the infections any time. These especial individuals are called infected individuals *I*(*t*). The individuals who are identified with coronavirus infections or COVID-19 and they are isolated to a separate place/ state for treatment. These individuals are described in the compartmental model as isolated individuals compartment and these individuals are denoted by *I*_*s*_ (*t*). The removed individuals *R*(*t*) are those who have recovered and thus have the immunity or have died from the disease and thus cannot contribute to further disease transmission. Figure 1 shows the transmission mechanisms of the novel coronavirus disease COVID-19.

Taking the above diagram into consideration, we formulate a five compartmental model with control in terms of a set of nonlinear ordinary differential equations (NODEs) of the following form:

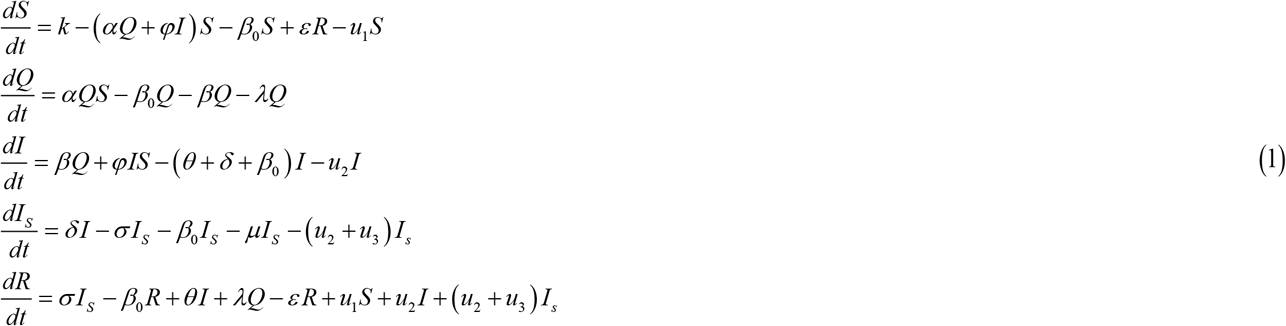

In the developed model, we have considered three control measures (*u*_1_, *u*_2_, *u*_3_) : (i) before infection, educational campaign among mass people so that the human to human transmission of coronavirus-2 infections can be prevented and (ii) after infection, (a) maintenance of social distancing which has been considered only the fruitful and effective step ever now to control the transmission of this horrible disease (COVID-19) and (b) treatment of the patients on the basis of the symptoms to minimize their sufferings. So that, *u*_1_ (*t*), *u*_2_ (*t*) and *u*_3_ (*t*) denotes the educational campaign, social distancing and treatment control respectively.

Model (1) is an optimal control model and the set of control measures (*u*_1_ (*t*), *u*_2_ (*t*),*u*_3_ (*t*))∈ *U* is Lebesgue measurable, where

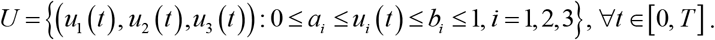

After considering these three control variables, we write the cost functional in the following form,

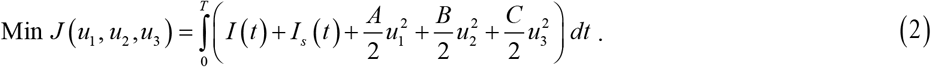

where *A, B* and *C* are the weight parameters or balancing parameters of the cost functional.

We can reformulate model (1) as the standard optimal control problem with respect to the cost functional (2) as

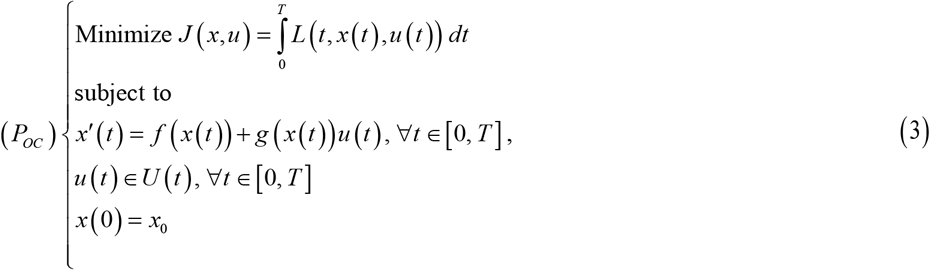

where, 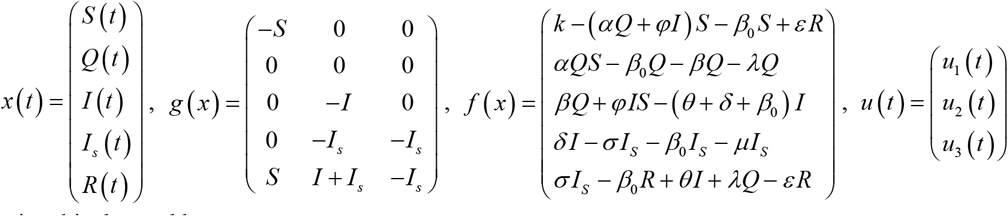 and the integrand of the cost functional is denoted by

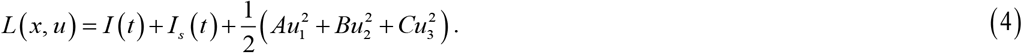

## 4. Characterization of the Optimal Control

We apply Pontryagin’s Maximum Principle to the Hamiltonian (H) in order to obtain the necessary conditions for this optimal control problem. Using Pontryagin’s Maximum Principle, the optimal control terms (such as educational campaign, social distancing and treatment) the standard Hamiltonian function H with respect to (*u*_1_, *u*_2_, *u*_3_) can be defined as follows:

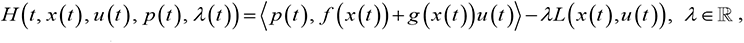

where 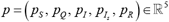 denotes the adjoint variables.

Suppose that the pair (*x*\*,*u*\*) is the optimal solution of the above optimal control problem. Then, the maximum principle states the existence of a scalar parameter *λ*_0_ ≥ 0, an absolutely continuous function *p*(*t*), such that the following conditions are held:

i. max{| *p*(*t*) |: *t* ∈[0, *T*]}+ *λ*_0_ > 0;
ii. -*p*′(*t*) = *λL*_*x*_ [*t*] − ⟨*p*[*t*], *f*_*x*_ [*t*] + *g*_*x*_ [*t*]*u*\* (*t*) ⟩;
iii. *p*(*t*) = (0, 0, 0, 0, 0) ;
iv. 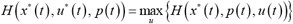, where *a*_1_ ≤ *u*_1_ ≤ *b*_1_, *a*_2_ ≤ *u*_2_ ≤ *b*_2_, *a*_3_ ≤ *u*_3_ ≤ *b*_3_,

where time argument [*t*] denotes the evaluation along with the optimal solution.

Then, from equation (ii) adjoint equations in normal form (i.e. *λ* = 1) are explicitly given by

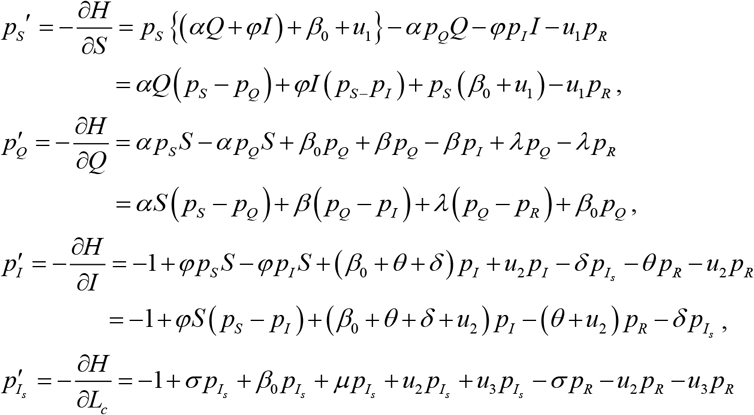

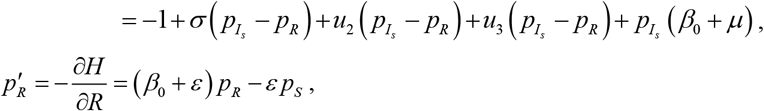

with transversality condition *p*_*i*_ (*T*) = 0, *i* = 1, 2, 3, 4 and 5.

## 5. Existence of the Optimal Control

In the present model, (*x*\*,*u*\*) is the optimal pair where *x*\* denotes the state variables and *u*\* represents control variables. So, in order to prove the existence of the optimal control, we have to show the existence of the state as well as the existence of the control variables.

### a. Existence of the State Variable

The state equation (1) with the initial condition can be written in the following form as

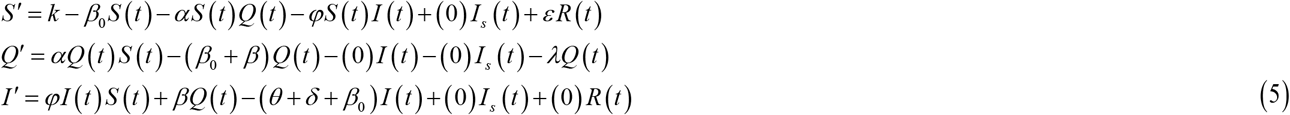

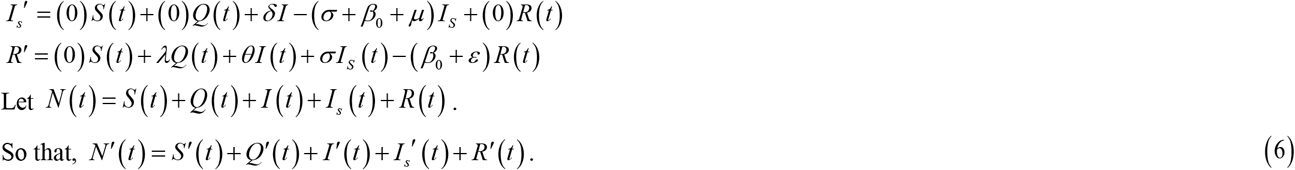

Now from equation (5) and (6), we can write

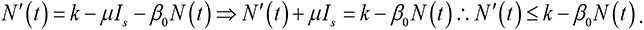

Here, *N*(*t*) is the total population.

So, we have 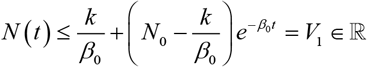 and 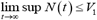, which gives *S*(*t*), *Q*(*t*), *I*(*t*), *I*_*s*_ (*t*), *R*(*t*) ≤ *V*_1_ as *t* →∞.

Then, we can rewrite equation (5) in the following form:

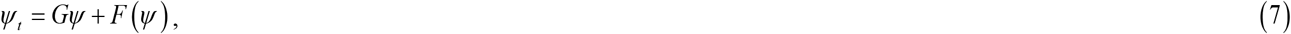

where 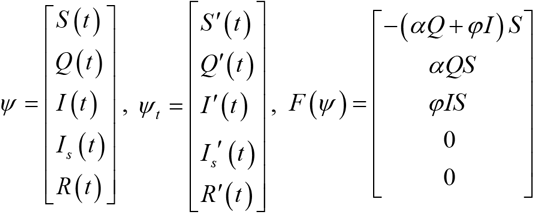 and 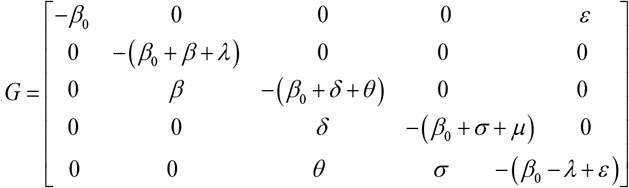.

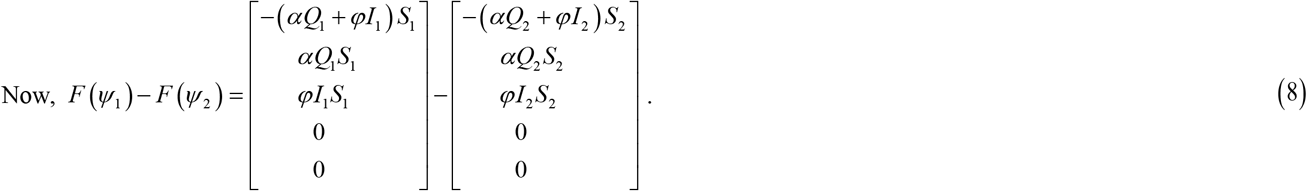

Equation (7) is a non-linear form with a bounded co-efficient.

We set *D*(*ψ*) = *ψ* _*t*_ = *G ψ* + *F*(*ψ*).

For the existence of optimal control and optimality system the boundedness of solution of the system for finite time is needed and we assume for *u* ∈*U*, there exists a bounded solution.

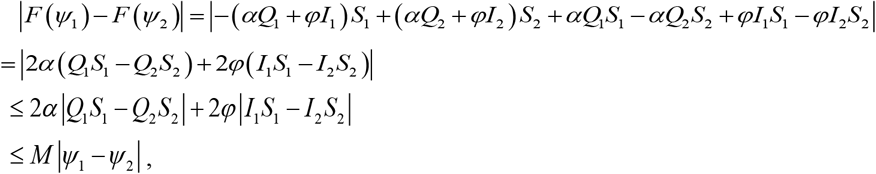

where, *M* is a constant.

Also, we get |*D*(*ψ*_1_) – *D*(*ψ*_2_)| ≤ ‖*B*‖ | *ψ*_1_ – *ψ*_2_ | + *M* | *ψ*_1_ – *ψ*_2_ | ≤ *V*|*ψ*_1_ – *ψ*_2_ |, where, *V* = max (*M*, ‖*B*‖) < ∞.

Thus, it follows that the function *D* is uniformly Lipschitz continuous. From the definition of the control *U*(*t*) and the restriction on *S, Q, I, I*_*s*_, and *R* ≥ 0, we see that a solution of the system (7) exists.

### b. Existence of the Control Variable

Now, by applying Pontryagin’s Maximum Principle [22] we have the following theorem and proving Theorem 1, we show the existence of controls.

**Theorem 1:** There exists optimal control (*u*_1_*,*u*_2_*,*u*_3_*) that minimizes the performance index *J*(*x, u*) over *U* given by

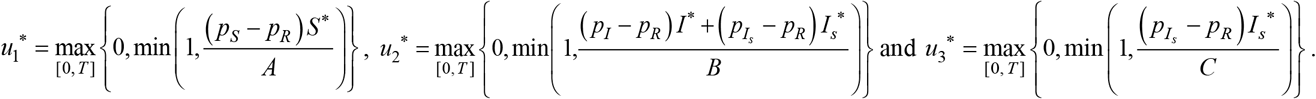

**Proof:** By optimality conditions, we have

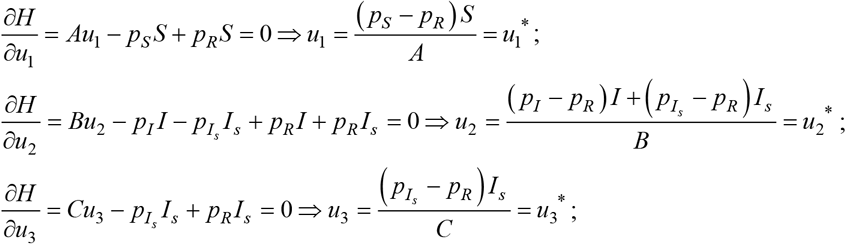

According to the property of *U*, the three controls (*u*_1_*, *u*_2_*, *u*_3_*) are bounded with upper bound 1and lower bound 0. Therefore,

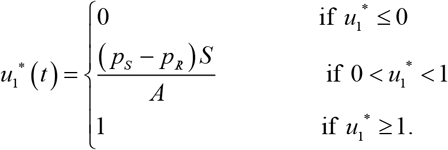

This can be written in compact form as

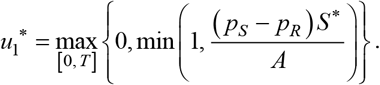

Similarly,

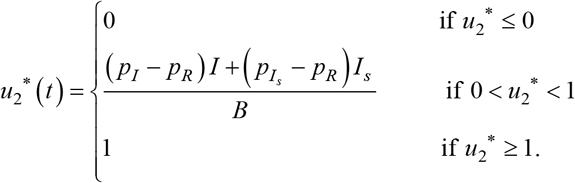

In the same way, this can be written in compact form as

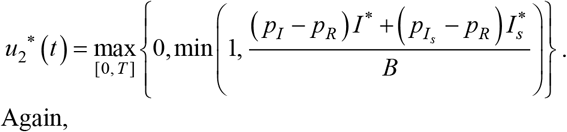

Again,

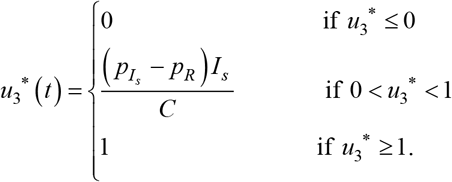

which can be written in compact form as

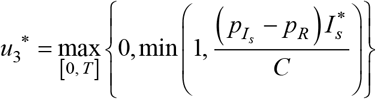

Thus, we get optimal solutions as

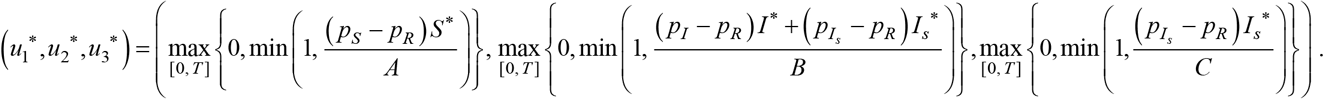

## 6. Numerical Results and Discussion

In this section, we have performed numerical simulations of optimal control model (3) using Open-OCL [16] written in MATLAB programming. In order to carry out numerical solutions of the model, we use a set of rational parameter values which are shown in Table 1. We simulate the model in two different ways. Firstly, we run the program of the model (1) without using any control variables and the simulated graph is shown in Figure 5. Furthermore, we perform the simulation of model (3) with control variables (such as educational campaign, social distancing and treatment). In this case, the results for each compartment are presented in Figures 6-9. Graphical outcomes are executed using the initial values: *S* = 10, 000, *Q* = 6, 000, *I* = 70, *I*_*s*_ = 3000, *R* = 30.

**Table 1:** Parameter specifications of model (3)

| Descriptions | Parameters | Values |
| --- | --- | --- |
| Source rate of susceptible individuals | $k$ | 19100 |
| Natural mortality rate | $\beta_0$ | 0.005 |
| Disease transmission rate | $\varphi$ | 0.3 |
| Quarantined rate | $\alpha$ | 0.6 |
| Coronavirus (CoV) relapse rate | $\varepsilon$ | 0.01 |
| Infection rate | $\beta$ | 0.8 |
| Recovery rate of quarantined individuals | $\lambda$ | 0.1 |
| Spontaneous recovery rate | $\theta$ | 0.7 |
| Isolation rate | $\delta$ | 0.1 |
| Cure rate from COVID 19 | $\sigma$ | 0.1 |
| Disease induced death rate | $\mu$ | 0.1 |

**Figure 4.**
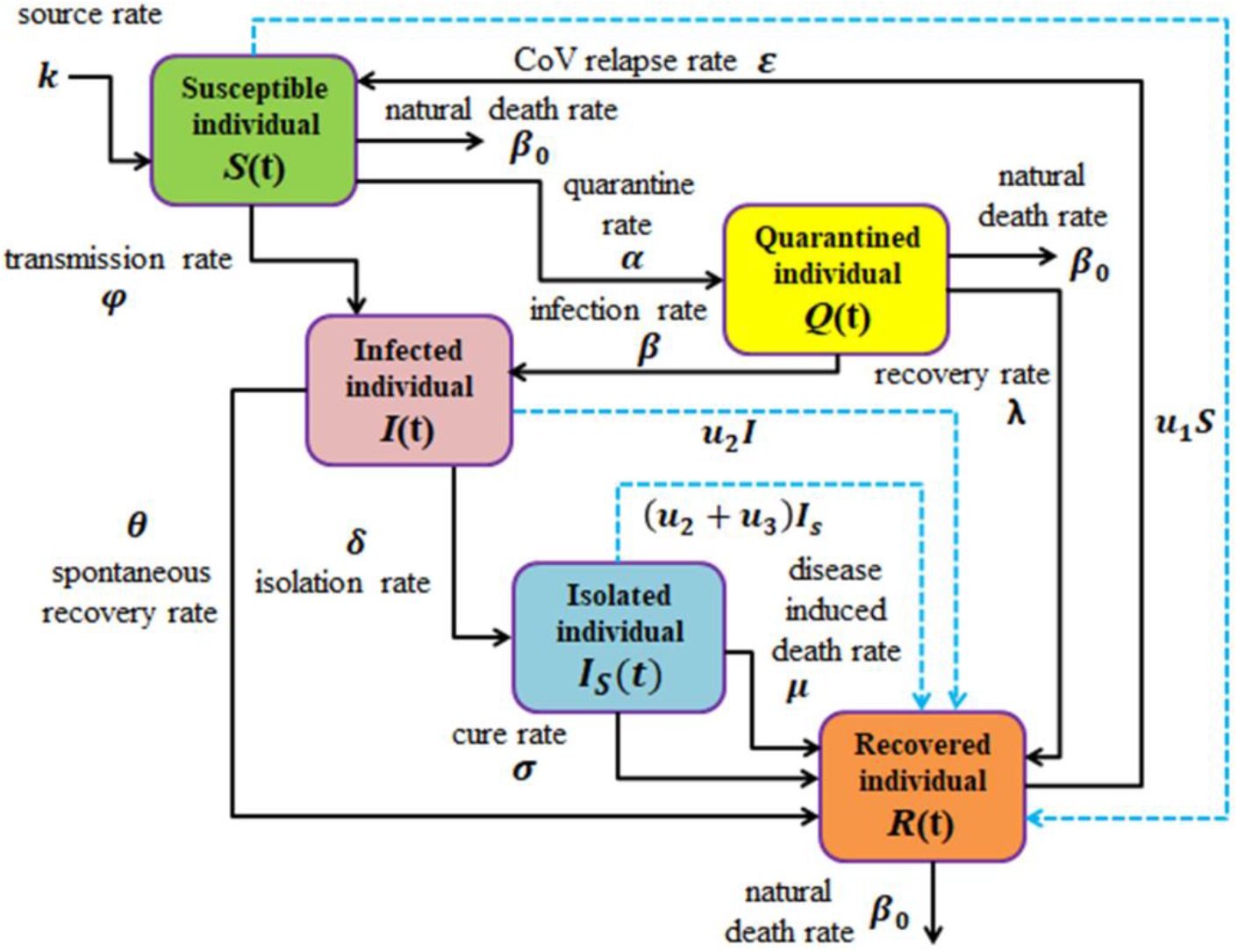
Flow chart of SIRS type compartmental model of SARS coronavirus-2 disease (COVID 19) with optimal control.

**Figure 5.**
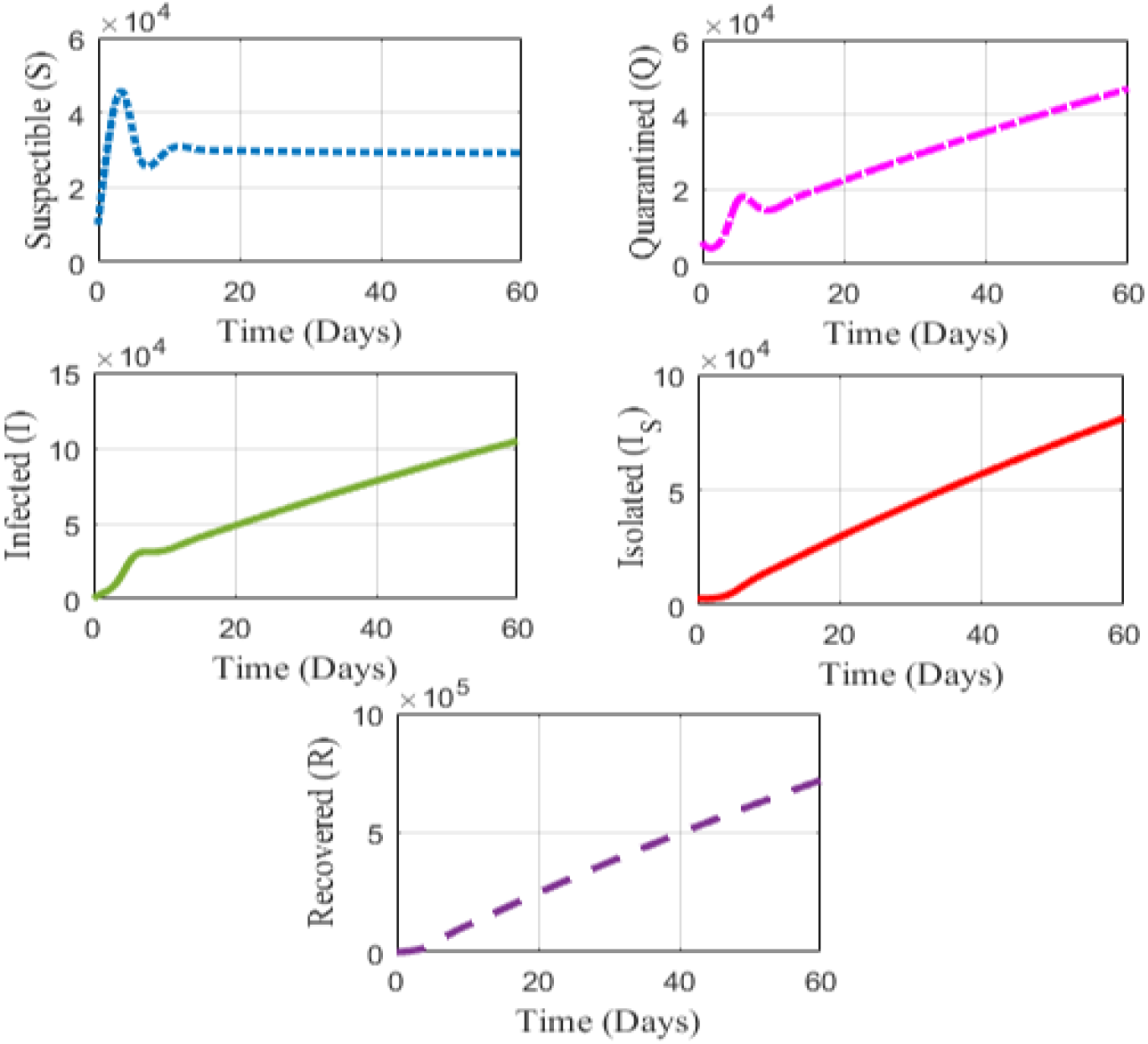
State trajectories of the compartmental model (1) when no control measures are employed (i.e. *u*_1_ = 0, *u*_2_ = 0, *u*_3_ = 0).

**Figure 6.**
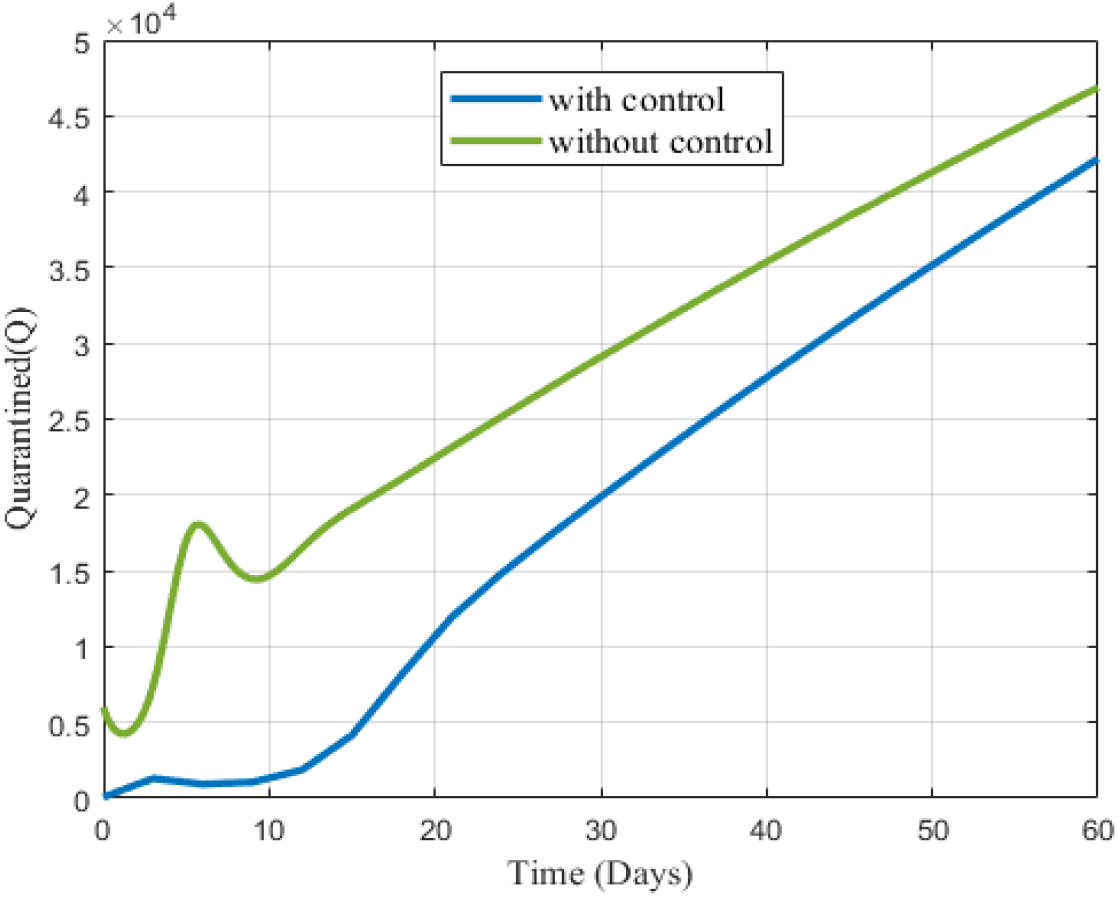
State trajectories of the quarantined individuals when educational campaign control (*u*_1_) and social distancing control (*u*_2_).

We have performed the whole simulations of the compartmental model (3) for both with and without control within 60 days timeline. For solving the model (3) with control, we simulate the optimal systems when all the three control variables (such as educational campaign, social distancing and treatment control) for coronavirus-2 disease (COVID 19) are employed. This time, we first run the program for the quarantined individuals for both cases (i.e. with and without control). Hence, the simulated outcome is illustrated in Figure 6. Basically, we adopt this in order to see the comparison of the different individuals (quarantined, infected, isolated and recovered individuals) for with and without control in the same window. We also run the program of the infected, isolated and recovered individuals for both with and without control separately. The simulated results are represented in Figures 7-9 respectively.

**Figure 7.**
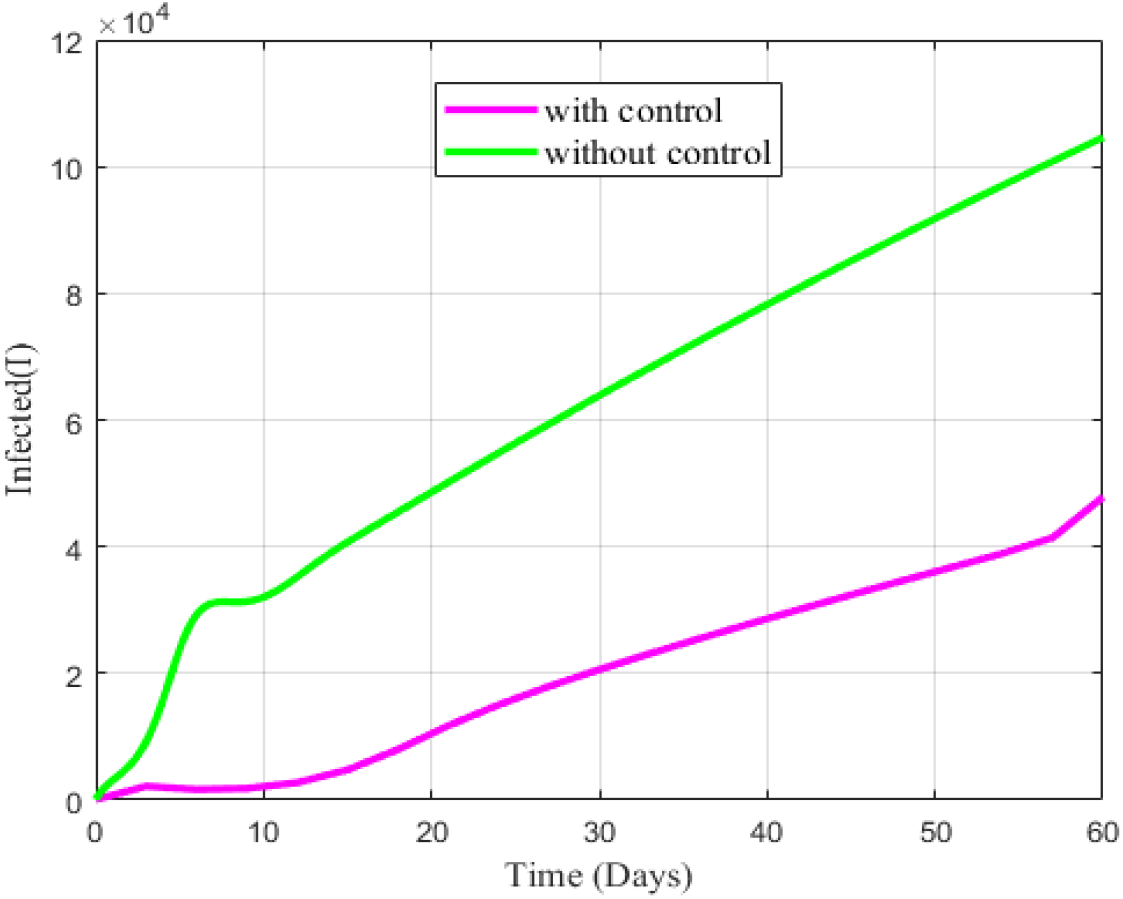
State trajectories of the infected individuals when educational campaign control (*u*_1_), social distancing control (*u*_2_) and treatment control (*u*_3_) are employed.

Figure 5 shows that the number of quarantined and infected individuals increases for first two days but probably in day three, it slightly reduces. From day five, the number of quarantined and infected individuals starts to grow whereas the number of isolated and recovered individuals soars from the initial state. However, it is noticeable that the number of susceptible individuals rises up for the first two days but for the next two days it decreases abruptly. Then it remains steady. This means that as the quarantined and infected individuals increase, the more individuals become isolated to get medical treatment as well as maintenance of social distancing, the more individuals get recovered.

Figure 6 illustrates the influences of educational campaign and social distancing control on the quarantined individuals for 60 days timeline. Here, it has been noticed that both the three control measures significantly control the number of quarantined individuals. The effects of the three controls are obvious in the figure. The number of quarantined individuals rises up when no control is employed but with the application of controls though it shows a slight decrease in days 1-3 and 9-10, it sharply plunges for the other time interval.

Figure 7 shows the influences of educational campaign, social distancing and treatment control on the infected individuals for 60 days timeline. From the figure, it has been observed that the number of infected individuals increases in the absence of control but when all three controls are employed, it shows a decreasing effect for the first eleven days and then gradually goes up. The reason behind the rise may be that the influence of educational campaign and treatment are not enough effective on infected individuals as no specific treatment for COVID-19 exists.

Figure 8 demonstrates the effects of three control measures such as educational campaign, social distancing and treatment control on the number of isolated individuals for 60 days timeline. In this case, it has been seen that the three control measures remarkably influences the isolated individuals. The isolated individual goes up with time without controls whereas it initially increases but gradually goes down for the first ten days and then it soars for the rest timeline with the application of the three controls.

**Figure 8.**
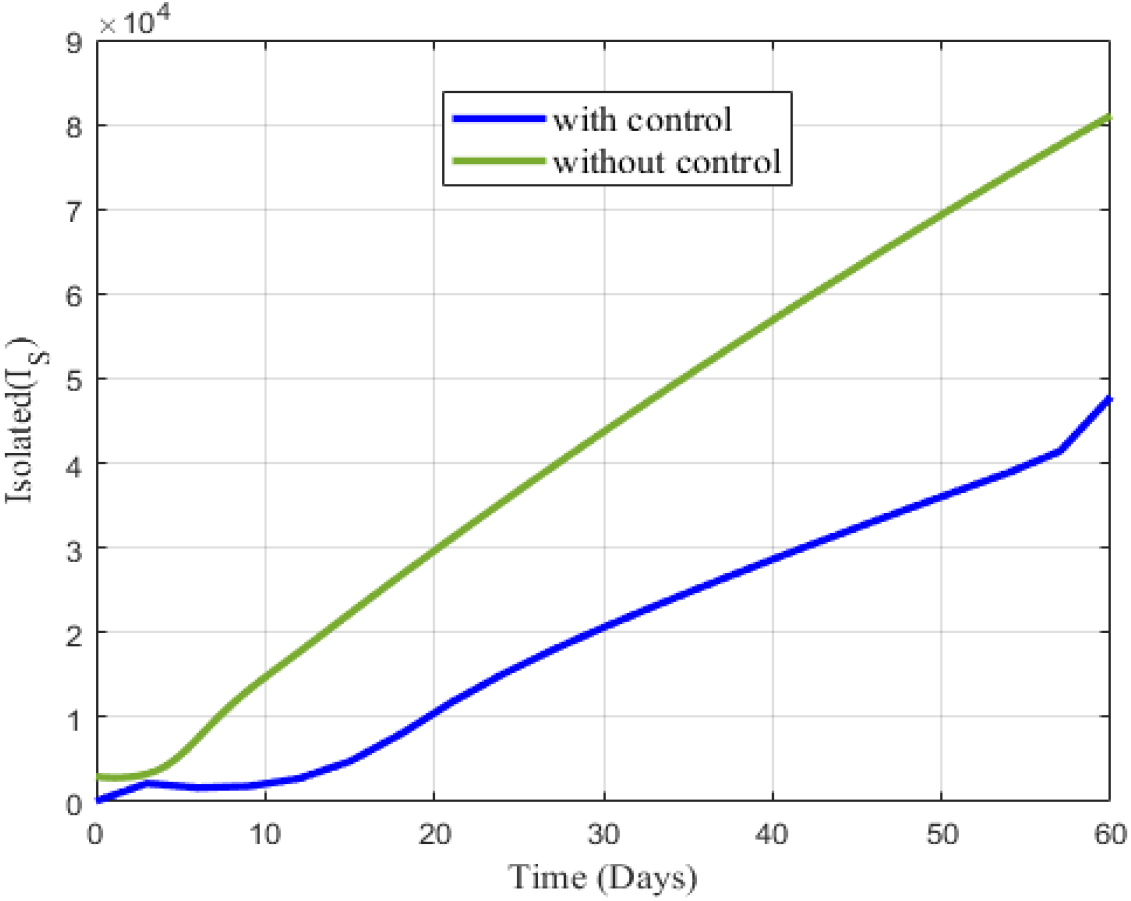
State trajectories of the isolated individuals when educational campaign control (*u*_1_), social distancing control (*u*_2_) and treatment control (*u*_3_) are activated.

Figure 9 depicts the effects of three control measures such as educational campaign, social distancing and treatment control on the number of recovered individuals for 60 days timeline. In this case, it is noticeable that the three control measures have significant influence on the recovered individuals. In the absence of control the number of recovered individuals is nearly zero for first ten days, then it shows a slight increase up to twenty days and finally it goes up. But this increase is lower than the expectation and also it is too small compared to the number of infected and isolated individuals. Hence to get the desired result, three controls have been employed and the recovered individuals swiftly booms for the implementation of educational campaign and social distancing as a prevention and treatment as cure of the disease COVID 19.

**Figure 9.**
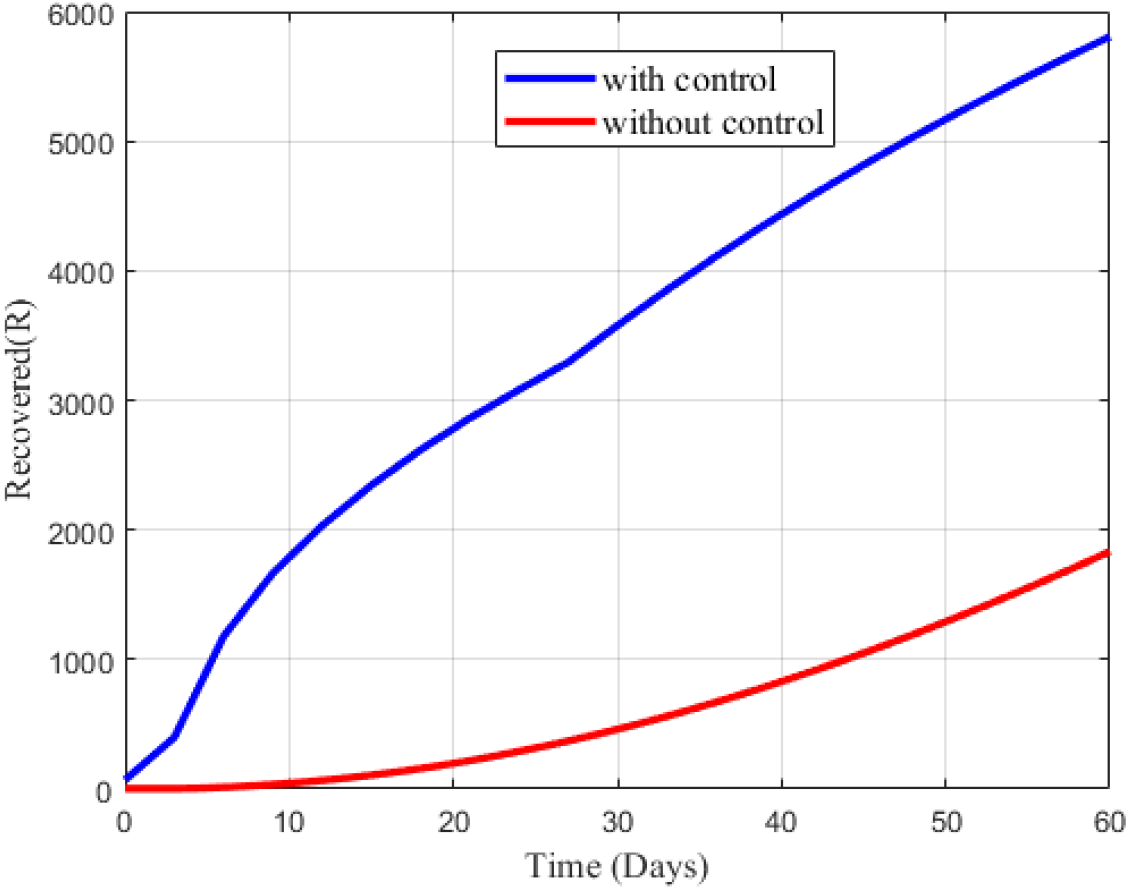
State trajectories of the quarantined individuals when educational campaign control (*u*_1_), social distancing control (*u*_2_) and treatment control (*u*_3_) are engaged.

## 7. Conclusions

COVID-19 is a highly contagious pandemic disease which is threatening the entire world with its devastating character. It is assumed that if the present trend of COVID-19 outbreak continues, then obviously humanity could face its most devastating pandemic. As there is no proper treatment with drugs or vaccines until to date, social distancing and educational campaign are the only ways to prevent and control people from being affected. Such a control strategy via mathematical modeling is discussed in the present study. The model is formulated considering three control variables by using the most well-known Pontryagin’s Maximum Principle. Numerical simulations have been performed to illustrate the analytic results. After investigation, it is observed that social distancing and educational campaign as prevention and treatment as cure are much more effective for the reduction of quarantined, infected and isolated individuals and at the same time the recovered individuals show a remarkable ascendancy. Since no antiviral medication (vaccination / natural health products) is authorized to treat or protect against COVID-19, implementation of educational campaign, social distancing and developing human immune system may be the best ways for preventing the disease before it goes out of control. Therefore, it is high time we took effective and strict measures to combat against this most devastating disease.

## Data Availability

The data used to support the findings of this study are included within the article.

## Acknowledgements

The authors greatly acknowledges the partial financial support provided by the Ministry of Science and Technology, Government of the People’s Republic of Bangladesh under special allocation in 2019-2020 with the research grant **Ref. No**.**-39**.**00**.**0000**.**009**.**06**.**024**.**19-12/410(EAS)**. Supports with Ref.: **17-392 RG/MATHS/AS_I– FR3240297753** funded by TWAS, Italy and Ref. no.-**6(74) UGC/ST/Physical-17/2017/3169** funded by the UGC, Bangladesh are also acknowledged.

## Data Availability

The data used to support the findings of this study are included within the article.

## Authors’ Contributions

This research is a group work carried out in collaboration among all authors. Author MHAB designed the study, performed the conceptualization and methodological analysis and model formulation of the first draft of the manuscript. Authors MSK and MRK analyzed the model analytically and wrote some literature of the study. Author AKP wrote the programming codes and performed some part of the computational analysis. Authors MAI and SAS contributed to literature searches and calculated the real data to estimate the parameters, UG verified the parameters and checked the literature. All authors have read and agreed to the publish the final version of the manuscript.

## References

1. Biswas, M.H.A., Paiva L.T. and de Pinho, M.D.R (2014). A SEIR Model for Control of Infectious Diseases with Constraints. Mathematical Biosciences and Engineering, 11(4), 761–784.

2. Biswas, M.H.A. (2014). On the Evaluation of AIDS/HIV Treatment: An optimal Control Approach. Current HIV Research, 12(1), 1–12.

3. Biswas, M.H.A. (2013). Necessary Conditions for Optimal Control Problems with State Constraints: Theory and Applications. PhD Thesis, Department of Electrical and Computer Engineering, Faculty of Engineering, University of Porto, Portugal.

4. Biswas, M.H.A. (2012). AIDS epidemic worldwide and the millennium development strategies: A light for lives. HIV and AIDS Review, 11(4), 87–94.

5. Biswas, M.H.A. (2014). Optimal Control of Nipah Virus (NiV) Infections: A Bangladesh Scenario. Journal of Pure and Applied Mathematics: Advances and Applications, 12(1), 77–104.

6. Biswas, M.H.A. (2013). On the Immunotherapy of HIV Infections via Optimal Control with Constraint. Proceedings of the 18th International Mathematics Conference, Dhaka, 20-22 March 2014, 51–54.

7. Biswas, M.H.A. (2012). Optimal Chemotherapeutic Strategy for HIV Infections-State Constrained Case. Proceedings of the 1st PhD Students Conference in Electrical and Computer Engineering, Department of Electrical and Computer Engineering, Faculty of Engineering, University of Porto, Portugal, 28-29 June 2012.

8. Biswas, M.H.A. (2012). Model and Control Strategy of the Deadly Nipah Virus (NiV) Infections in Bangladesh. Research &Reviews in Biosciences, 6(12), 370–377.

9. Biswas, M.H.A., Haque, M.M. and Mallick U.K. (2019). Optimal Control Strategy for the Immunotherapeutic Treatment of HIV Infection with State Constraint. Optimal Control, Applications and Methods, 40(3), 1–12.

10. COVID-19 Coronavirus Pandemic, Worldometer. Retrieved from https://www.worldometers.info/coronavirus/. (Accessed on 2 April)

11. COVID-19 Coronavirus, China, Worldometer. Retrieved from https://www.worldometers.info/coronavirus/country/china/. (Accessed on 2 April)

12. COVID-19 Situation in the WHO South East-Asia Region, World Health Organization (WHO), South East Asia. Retrieved from https://www.who.int/southeastasia/outbreaks-and-emergencies/novel-coronavirus-2019. (Accessed on 2 April)

13. Chen, X. and Yu, B. (2020). First Two Months of the 2019 Coronavirus Disease (COVID-19) Epidemic in China: Real Time Surveillance and Evaluation with a Second Derivative Model, Global Health Research and Policy, 5, 7. https://doi.org/10.1186/s41256-020-00137-4

14. Chen, T.M., Rui, J., Wang, Q., Zhao, Z., Cui, J. and Yin, L. (2020). A Mathematical Model for Simulating the Phase-Based Transmissibility of A Novel Coronavirus, Infectious Diseases of Poverty, 9, 24. https://doi.org/10.1186/s40249-020-00640-3

15. Fong, S. J., Li, G., Dey, N., Crespo, R. G. and Viedma, E.H. (2020). Finding an Accurate Early Forecasting Model from Small Dataset: A Case of 2019-nCoV Novel Coronavirus Outbreak, International Journal of Interactive Multimedia and Artificial Intelligence, 6(1), 132–140.

16. Koenemann, J., Licitra, G., Alp, M. and Diehl, M. (2019). ‘OpenOCL-Open Optimal Control Library’ Robotics Science and Systems, workshop submission, extended abstract, June 2019.

17. Kucharski, A.J., Russell, T. W., Diamond, C., Liu, Y., Edmunds, J., Funk, S. and Eggo M.E. (2020). Early Dynamics of Transmission and Control Of COVID-19: A Mathematical Modelling Study, The Lancet Infectious Diseases, 20, 30144–4.

18. Khan, M. A. and Atangana, A. (2020). Modeling the Dynamics of Novel Coronavirus (2019-nCov) with Fractional Derivative, Alexandria Engineering Journal. https://doi.org/10.1016/j.aej.2020.02.033

19. Khatun, M.S. and Biswas, M.H.A. (2020). Optimal control strategies for preventing hepatitis B infection and reducing chronic liver cirrhosis incidence, Infectious Disease Modelling, 5(2020), 91–110.

20. Khatun, M.S. and Biswas, M.H.A. (2019). Modeling the Effect of Adoptive T cell Therapy for the Treatment of Leukemia,Computational and Mathematical Method, doi: 10.1002/cmm4.1069.

21. Lin Q., Zhao, S., Gao, D., Lou, Y., Wang, S., Musa, S.S., Wang, M. H., Cai, Y., Wang, W., Yang, L. and Daihai, H. (2020). A Conceptual Model for the Coronavirus Disease 2019 (COVID-19) Outbreak in Wuhan, China with Individual Reaction and Governmental Action, International Journal of Infectious Diseases, 93, 211–216.

22. Lenhart, S. and Workman, J.T. (2007). “Optimal control applied to biological models”, Chapman & Hall, New York.

23. Sookaromdee, P. and Wiwanitkit, V. (2020). Imported Cases of 2019-Novel Coronavirus (2019-Ncov) Infections in Thailand: Mathematical Modelling of The Outbreak, Asian Pacific Journal of Tropical Medicine, 13(3), 139–140.

24. Severe Acute Respiratory Syndrome, World Health Organization. Retrieved from https://www.who.int/ith/diseases/sars/en/. (Accessed on 2 April)

25. Wu, J., Leung, K. and Leung, G. M. (2020). Nowcasting And Forecasting the Potential Domestic and International Spread of the 2019-Ncov Outbreak Originating in Wuhan, China: A Modelling Study, Lancet, 395(10225), 689–697.

26. Xu, X., Chen, P., Wang, J., Feng, J., Zhou, H., Li, X., Zhong, W. and Hao, P. (2020). Evolution of the Novel Coronavirus from the Ongoing Wuhan Outbreak and Modeling of Its Spike Protein for Risk of Human Transmission. Science China Life Sciences, 63, 457–460.

27. Zhao, S., Salihu, S., Qianying, L., Jinjun ., Yang, G., Wang, W., Lou Y., Yang L., Gao D., Daihai, H. and Maggie, H. W. (2020). Estimating the Unreported Number of Novel Coronavirus (2019-nCoV) Cases in China in the First Half of January 2020: A Data-Driven Modelling Analysis of the Early Outbreak, Journal of Clinical Medicine, 9, 388. https://doi.org/10.3390/jcm9020388

28. Zhao, S., Lin Q., Ran, J., Musa, S.S., Yang, G., Wang, W., Lou, Y., Gao, D., Yang, L., He, D. and Wang, M. H. (2020). Preliminary Estimation of the Basic Reproduction Number of Novel Coronavirus (2019-nCoV) in China, from 2019 to 2020: A Data-Driven Analysis in the Early Phase of the Outbreak, International Journal of Infectious Diseases, 92, 214–217.

29. Zhang, J., Weili, W., Zhao, X., and Zhang, W. (2020). Recommended Psychological Crisis Intervention Response to the 2019 Novel Coronavirus Pneumonia Outbreak in China: A Model of West China Hospital, Precision Clinical Medicine, 3(1), 3–8.

30. Zhang, S., Diao, M., Wenbo, Y., Pei L., Lin, Z. and Chen D. (2020). Estimation of the Reproductive Number of Novel Coronavirus (COVID-19) and the Probable Outbreak Size on the Diamond Princess Cruise Ship: A Data-driven Analysis,International Journal of Infectious Diseases, 93, 201–214.

31. Zhuang, Z., Zhao, S., Lin. Q., Cao, P., Lou, Y., Yang, L. and Daihai, H. (2020). “Preliminary Estimation of the Novel Coronavirus Disease (COVID-19) Cases in Iran: A Modelling Analysis Based on Overseas Cases and Air Travel Data” https://doi.org/10.1101/2020.03.02.20030312

